# Precautions are Needed for COVID-19 Patients with Coinfection of Common Respiratory Pathogens

**DOI:** 10.1101/2020.02.29.20027698

**Authors:** Quan-sheng Xing, Guo-ju Li, Yu-han Xing, Ting Chen, Wen-jie Li, Wei Ni, Kai Deng, Ru-qin Gao, Chang-zheng Chen, Yang Gao, Qiang Li, Gui-ling Yu, Jian-ning Tong, Wei Li, Gui-liang Hao, Yue Sun, Ai Zhang, Qin Wu, Zi-pu Li, Si-lin Pan

**Affiliations:** Qingdao Women and Children’s Hospital, Qingdao University, Qingdao City, Shandong Province, China; Department of Paediatrics, Faculty of Medicine, The Chinese University of Hong Kong, Hong Kong SAR, China; Department of Ophthalmology, Renmin Hospital of Wuhan University, Wuhan City, Hubei Province, China; Qingdao Chest Hospital, Qingdao City, Shandong Province, China; Qingdao Municipal Centre for Disease Control and Prevention, Qingdao Institute of Prevention Medicine, Qingdao City, Shandong Province, China

**Author notes:** Contributed equally (Quan-sheng Xing, Guo-ju Li, Yu-han Xing, Ting Chen, Wen-jie Li, Wei Ni, Kai Deng, Ru-qin Gao, Chang-zheng Chen). **Correspondence** Quansheng Xing, MD, PhD: Qingdao Women and Children’s Hospital, Qingdao University, Qingdao, Shandong Province, China, No.6 Tongfu Road, 266000, Qingdao, China.

## Abstract

**Background:** With the ongoing outbreak of Coronavirus Disease 2019 (COVID-19), infected patients within and beyond the epidemic area, Wuhan, China, showed different epidemiological and clinical characteristics. There is a paucity of data concerning coinfection with other common respiratory pathogens in COVID-19 patients outside of Wuhan.

**Methods:** We conducted a double-centre study recruiting 68 patients with severe acute respiratory coronavirus 2 (SARS-CoV-2) infection confirmed by nucleic acid testing in Qingdao and Wuhan from January 17 to February 16, 2020. Indirect immunofluorescence was performed to detect the specific IgM antibody against common respiratory pathogens in collected acute phase serum.

**Results:** Of the 68 patients with SARS-CoV-2 infection, 30 (44.12%) were from Qingdao. The median age of Qingdao and Wuhan patients were 50 (IQR: 37-59) and 31 (IQR: 28-38) years, respectively, and the majority of patients were female in Qingdao (60.00%) and Wuhan (55.26%). Among COVID-19 patients in Qingdao, 24 (80.00%) of them had IgM antibodies against at least one respiratory pathogen, whereas only one (2.63%) of the patients in Wuhan had positive results for serum IgM antibody detection (*P*<0.0001). The most common respiratory pathogens detected in Qingdao COVID-19 patients were influenza virus A (60.00%) and influenza virus B (53.33%), followed by *mycoplasma pneumoniae* (23.33%) and *legionella pneumophila* (20.00%). While the pattern for coinfection in patients with community-acquired pneumonia in Qingdao was quite different, with a positive rate of only 20.90%.

**Interpretation:** We reported a large proportion of COVID-19 patients with coinfection of seasonal respiratory pathogens in Qingdao, northeast China, which differed greatly from the patients in Wuhan, central China. Precautions are needed when dealing with COVID-19 patients beyond the epidemic centre who have coinfection with other respiratory pathogens. We highly recommend adding SARS-CoV-2 to routine diagnostic testing in capable hospitals to prevent misdetection of the virus.

## Introduction

At the beginning of December 2019, a cluster of “pneumonia of unknown aetiology” emerged in Wuhan, Hubei Province, China. The disease has soon developed into an outbreak posing a pandemic threat. Since no causative pathogen was identified at the onset of the disease, it was once called “Wuhan pneumonia” by the health officials and the public. On December 31, 2019, a total of 27 cases were reported; meanwhile, a rapid response team led by the Chinese Centre for Disease Control and Prevention (China CDC) was formed to conduct detailed epidemiologic and aetiologic investigations in Wuhan.^1-3^ After ruling out common respiratory pathogens such as influenza, avian influenza, respiratory syncytial virus (RSV), adenovirus (ADV), respiratory severe acute respiratory syndrome coronavirus (SARS-CoV) and Middle East respiratory syndrome coronavirus (MERS-CoV), etc., a novel coronavirus was confirmed as the aetiological agent on January 7, 2020 and related disease was termed as Coronavirus Disease 2019 (COVID-19) by the World Health Organisation (WHO).^4^ Subsequent full-length genome sequencing analysis indicated that the newly discovered virus, now known as severe acute respiratory coronavirus 2 (SARS-CoV-2), belongs to a distinct beta-coronavirus genus of probable bat origin.^5,6^ Thus far, the epidemic has swept through the nation. At the time of writing, the number of confirmed COVID-19 cases in China has increased to 79,968, causing 2,873 deaths.^7^ The fast-growing outbreak has affected over fifty countries and regions beyond China, constituting a public health emergency of international concern.^8^

Strikingly, we found a large proportion of coinfection with other seasonal respiratory pathogens in COVID-19 patients admitted to hospitals in Qingdao, Shandong Province, northeast China, which differed from those of the primary infected patients from the epidemic centre, Wuhan, central China.^3,9^ To determine such between-region difference and to explore the underlying reasons, we conducted this double-centre study recruiting COVID-19 patients admitted in Qingdao and Wuhan hospitals. We hope our study will provide new insights into clinical diagnosis and prophylactic control of COVID-19 and to facilitate an in-depth understanding of the disease.

## Methods

### Study areas

We chose Qingdao and Wuhan as the study centres to investigate the difference in coinfection with common respiratory pathogens among COVID-19 patients. **Figure 1** showed the geographical location of two cities. As depicted, Qingdao is situated about 850 kilometres northeast of Wuhan.

**Figure 1.**
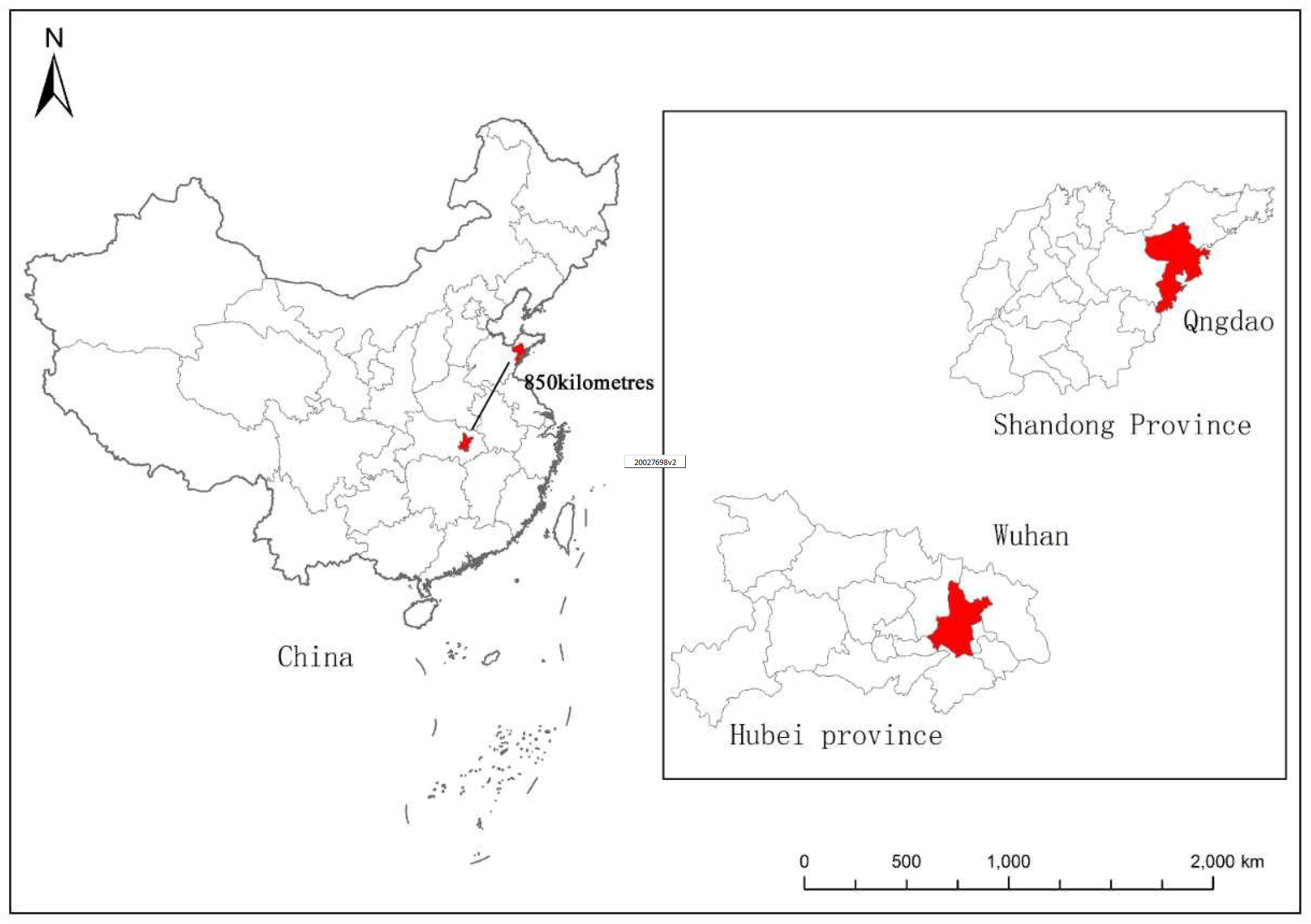
Geographical location of Qingdao and Wuhan, China

### Participants

We recruited patients with laboratory-confirmed COVID-19 who admitted to the 3 designated hospitals in Qingdao (Qingdao Women and Children’s Hospital, Affiliated Hospital of Qingdao University Medical College, and Qingdao Chest Hospital) and Renmin Hospital of Wuhan University from January 17 to February 16, 2020. Laboratory testing was done for all patients for the detection of SARS-CoV-2 and other respiratory pathogens. The epidemiological characteristics, clinical presentations, and laboratory findings were extracted from electronic medical records. Radiographic evidence included chest X-ray or computed tomography (CT). Laboratory assessments consisted of blood routine, blood biochemistry, liver function and C-reactive protein measurements. All patients had positive results of nucleic acid test of SARS-CoV-2 in respiratory tract specimens. The diagnosis of COVID-19 was made according to WHO interim guidance and the severity of disease was mild to moderate.^4^ Seven cases in Qingdao were not included in demographic and clinical analysis due to lack of related information. To test the reliability of pathogen detection method, we randomly selected 16 healthy blood donors and 14 asymptomatic individuals who came to the hospitals in Qingdao for health counselling as the group for quality control (healthy control group). In addition, 67 patients with community-acquired pneumonia (CAP) who admitted to the local hospitals in Qingdao at the same time were enrolled as a comparable group to COVID-19 patient group (CAP control group). Diagnosis of CAP was based on the latest clinical guideline.^10^

This study was approved by the Ethics Commission of each participating hospital (QFFLL-KY-2020-11) and written informed consent was obtained from involved patients prior to enrolment.

### Detection of SARS-CoV-2 in respiratory samples

Laboratory testing of SARS-CoV-2 was done in biosafety level 3 (P3) laboratory at individual hospital within 2 hours after sample collection; samples collected in Qingdao were also double-tested by Municipal Centre of Disease Control and Prevention of Qingdao. Throat swabs were obtained from all patients upon admission using standardised techniques in negative-pressure isolation rooms. After collection, throat swab was kept in 1.5 mL virus preservation solution for subsequent extraction of total RNA using a magnetic bead-based kit (HEALTH BioMed Co., Ltd, Ningbo, China). The presence of SARS-CoV-2 was detected by real-time fluorescence reverse-transcriptase-polymerase-chain reaction (RT-PCR). Both the commercial kits used in Qingdao (DAAN Gene Co., Ltd of Sun Yat-sen University, China) and Wuhan (GeneoDX Co., Ltd, Shanghai, China) were approved by the China Food and Drug Administration (CFDA). For samples tested in Qingdao, the PCR assay simultaneously amplified two target genes of SARS-CoV-2 included open reading frame 1ab (*ORF1ab*) and nucleocapsid protein (N) and the sequences were as follows: forward primer 5′-CCCTGTGGGTTTTACACTTAA-3′, reverse primer 5′-ACGATTGTGCATCAGCTGA-3′; and the probe 5′-FAM-CCGTCTGCGGTATGTGGAAAGGTTATGG-BHQ1-3′. Amplification was performed under the following conditions: incubation at 50 °C for 15 min and 95 °C for 15 min, followed by 45 cycles of denaturation at 95 °C for 15 sec, and extending and collecting fluorescence signal at 55 °C for 15 sec (7500 Real-Time PCR Systems, Applied Biosystems™, Thermo Fisher Scientific). A cycle threshold (Ct) value no more than 40 with evident amplification curve was considered as a positive test, and a Ct value over 40 was defined as a negative result. Procedures of viral detection done in Wuhan was described elsewhere.^11^

### Detection of serum IgM antibody against respiratory pathogens

Laboratories in Qingdao and Wuhan adopted a similar protocol for detection of IgM-specific antibodies against respiratory pathogens in collected acute phase serum. Indirect immunofluorescence (IIFA) was performed to detect serum IgM antibody by using a commercially available kit (samples collected from Qingdao patients: EUROIMMUN Medizinische Labor diagnostika AG, CN; samples collected from Wuhan patients: Vircell, S.L., Santa Fe’, Granada, Spain) according to manufacturer’s instructions.^12^ IIFA can detect 9 of the common respiratory pathogens including respiratory syncytial virus (RSV), adenovirus (ADV), influenza virus A (IFV-A), influenza virus B (IFV-B), parainfluenza virus (PIV), *mycoplasma pneumoniae* (MP), *chlamydia pneumoniae* (CP), *legionella pneumophila* (LP) and *Coxiella burnetii* (COX) in a single assay. Briefly, diluted serum solution (1:100 for detection of LP and 1:10 for other pathogens) was incubated on each well of the slide and stained with fluorescein-conjugated antibodies. The multi-well slide was then processed for fixing, staining, washing and drying steps, the entire well containing the stained specimen was examined under an epifluorescence microscope. RT-PCR nucleic acid test was also performed in respiratory specimens collected from Wuhan patients for detection of other 13 respiratory viruses including IFV-A, IFV-B, H1N1, H3N2, PIV, RSV, human metapneumovirus, SARS-CoV, rhinovirus, ADV, Bocavirus, MP and CP.

### Statistical analysis

Raw data were entered by two persons (double data entry) who were not aware of the arrangement of study groups. Continuous variables (non-normal distribution) were expressed as median with interquartile range (IQR) and compared with the Mann-Whitney U test; categorical variables were presented as number (%) and compared by χ^2^ test or Fisher’s exact test between Wuhan and Qingdao groups. A two-sided α of less than 0.05 was considered statistically significant. Statistical analyses were done using the SAS software, version 9.4.

## Results

By February 16, 2020, a total of 68 patients with laboratory-confirmed SARS-CoV-2 infection was included in the final analysis, among whom 30 were from Qingdao and 38 were from Wuhan (**Table 1**). The median age of Qingdao and Wuhan patients were 50 years (IQR: 37-59) and 31 years (IQR: 28-38), respectively. The median duration from symptom onset to hospitalisation was 2 days (IQR: 0-5) for patients in Qingdao and 4 days (IQR: 2-5) for that in Wuhan. The majority of patients admitted in Qingdao and Wuhan hospitals were female, representing a proportion of 60.00% and 55.26%, respectively. The most common symptoms at onset of COVID-19 were fever (66.67% *vs*. 57.89%) and cough (50.00% *vs*. 68.42%) in both Qingdao and Wuhan patients. Over one third of the Qingdao patients had underlying diseases (36.67%), a proportion much higher than that of Wuhan patients (2.63%). In comparison with Wuhan patients, Qingdao patients were older, had a higher respiratory rate (20 breaths/min [IQR: 19-21] *vs*. 19 breaths/min [IQR: 18-20]) and increased diastolic blood pressure (77 mmHg [IQR: 70-84] *vs*. 72 mmHg [IQR: 70-77]; all *P* values less than 0.05). On admission, white blood cell and platelet counts of Qingdao patients were higher than those of Wuhan patients (median white blood cell count: 5.11×10^9^/L [IQR: 4.15-6.10] *vs*. 4.17×10^9^/L [IQR: 3.50-5.60], *P=*0.0401; median platelet count: 237×10^9^/L [IQR: 169-265] *vs*. 166× 10^9^/L [IQR: 148-207], *P=*0.0043). The median total bilirubin level was 12.50 μmol/L (IQR: 10.50-15.50) in Qingdao patients, which was significantly higher than that in Wuhan patients (7.14 μmol/L [IQR: 5.70-9.70], *P=*0.0016).

**Table 1.** Characteristics of patients infected with SARS-CoV-2.

|  | **Qingdao (n=30)** | **Wuhan (n=38)** | ***P*-value** |
| --- | --- | --- | --- |
| Age, y | 50 (37-59) | 31 (28-38) | **0.0009** |
| Male | 12 (40.00) | 17 (44.74) | 0.6949 |
| In hospital day | 2 (0-5) | 4 (2-5) | 0.2350 |
| Fever | 20 (66.67) | 22 (57.89) | 0.4599 |
| Cough | 15 (50.00) | 26 (68.42) | 0.1232 |
| Dyspnea | 0 (0.00) | 3 (7.89) | 0.2493 |
| Myalgia | 5 (16.67) | 11 (28.95) | 0.2359 |
| Headache | 4 (13.33) | 3 (7.89) | 0.6909 |
| Pharyngalgia | 4 (13.33) | 6 (15.79) | 1.0000 |
| Fatigue | 8 (26.67) | 15 (39.47) | 0.2677 |
| Anorexia | 4 (13.33) | 10 (26.32) | 0.1886 |
| Diarrhea | 5 (16.67) | 2 (5.26) | 0.2272 |
| Expectoration | 8 (26.67) | 9 (23.68) | 0.7779 |
| Dizziness | 1 (3.33) | 2 (5.26) | 1.0000 |
| Chest congestion | 6 (20.00) | 2 (5.26) | 0.1256 |
| Heart rate, bpm | 79 (73-91) | 78 (76-81) | 0.3817 |
| Respiratory rate, min | 20 (19-21) | 19 (18-20) | **0.0033** |
| SBP, mmHg | 120 (114-133) | 120 (117-125) | 0.5596 |
| DBP, mmHg | 77 (70-84) | 72 (70-77) | **0.0413** |
| Temperature, °C | 36.9 (36.4-37.0) | 36.6 (36.5-36.8) | 0.0900 |
| Combined diseases | 11 (36.67) | 1 (2.63) | **0.0003** |
| Clustering of the symptoms, $\geq 4$ | 8 (26.67) | 13 (34.21) | 0.5038 |

The data are expressed as n (%) or median (IQR).

**Table 2.** Laboratory findings of patients infected with SARS-CoV-2.

|  | **Qingdao (n=30)** | **Wuhan (n=38)** | **P-value** |
| --- | --- | --- | --- |
| **Blood routine** |  |  |  |
| White blood cell count, $\times 10^9/L$ | 5.11 (4.15-6.10) | 4.17 (3.50-5.60) | **0.0401** |
| Neutrophils$\times 10^9/L$ | 3.17 (2.62-3.59) | 2.49 (1.89-3.57) | 0.0788 |
| Lymphocyte$\times 10^9/L$ | 1.26 (1.12-1.78) | 1.19 (1.00-1.64) | 0.3102 |
| Platelet$\times 10^9/L$ | 237 (169-265) | 166 (148-207) | **0.0043** |
| Hemoglobin, g/L | 130 (123-149) | 139 (128-148) | 0.2239 |
| **Blood biochemistry** |  |  |  |
| Albumin, g/L | 43.20 (38.80-44.40) | 42.05 (40.50-44.30) | 0.8012 |
| Alanine aminotransferase, U/L | 19.30 (12.00-26.00) | 19.00 (13.00-31.00) | 0.6213 |
| Aspartate<br>aminotransferase, U/L | 22.00 (19.00-31.00) | 21.00 (18.00-29.00) | 0.4217 |
| Total bilirubin, $\mu\text{mol/L}$ | 12.50 (10.50-15.50) | 7.14 (5.70-9.70) | **0.0016** |
| Serum creatinine, $\mu\text{mol/L}$ | 61.00 (43.00-73.00) | 60.00 (46.00-72.00) | 0.8134 |
| Glucose, mmol/L | 5.20 (4.71-8.40) | 4.91 (4.51-5.55) | 0.0823 |
| **Infection-related biomarkers** |  |  |  |
| C-reactive protein, mg/L,<br>(normal range: 0.0-10.0) |  |  |  |
| Increased | 14 (46.67) | 10 (27.03) | 0.0812 |

The data are expressed as n (%) or median (IQR).

All patients had respiratory specimens tested for specific IgM antibodies against IFV-A, IFV-B, RSV, ADV, PIV, MP, LP, CP and COX. Among the 30 patients admitted in Qingdao, 24 patients had IgM antibodies detected against at least one of the above-mentioned pathogens, and the overall positive rate was 80.00% (**Table 3**); whereas only one (2.63%) of the patients in Wuhan had positive results for respiratory pathogens. The most common respiratory viruses detected were IFV-A (60.00%) and IFV-B (53.33%), followed by MP (23.33%) and LP (20.00%).

**Table 3.** Coinfection of common respiratory pathogens in COVID-19 patients in Qingdao and Wuhan.

| Pathogens detected | Qingdao (n=30) | Wuhan (n=38) | *P*-value |
| --- | --- | --- | --- |
|  | No. (%) | No. (%) |  |
| IFV-A | 18 (60.00) | 0 (0.00) | **<0.0001** |
| IFV-B | 16 (53.33) | 0 (0.00) | **<0.0001** |
| MP | 7 (23.33) | 1 (2.63) | **0.0179** |
| LP | 6 (20.00) | 0 (0.00) | **0.0054** |
| RSV | 0 (0.00) | 1 (2.63) | 1.0000 |
| ADV | 0 (0.00) | 0 (0.00) | NA |
| PIV | 0 (0.00) | 0 (0.00) | NA |
| CP | 0 (0.00) | 0 (0.00) | NA |
| COX | 0 (0.00) | 0 (0.00) | NA |
| Total | 24 (80.00) | 1 (2.63) | **<0.0001** |

Abbreviations: influenza A virus (IFV-A), influenza B virus (IFV-B), *mycoplasma pneumoniae* (MP), *legionella pneumophila* (LP), respiratory syncytial virus (RSV), adenovirus (ADV), parainfluenza virus (PIV), *chlamydia pneumoniae* (CP) and *Coxiella burnetii* (COX).

Ages of healthy control group ranged from 20 to 55 years, with a median age of 40 years (IQR: 33-50), and 14 (46.67%) were men. Only 4 people (13.33%) of this group had specific-IgM antibody detected in their serum, suggesting asymptomatic infection with single virus (IFV-B). The total infection rate in healthy control was significantly lower than that in COVID-19 patients (*P*<0.0001, **Table 4**), and none of the individual in control group showed evidence of combined infection with two or more pathogens.

**Table 4.** Common respiratory pathogens detected in COVID-19 patients and healthy controls.

| **Pathogens detected** | **COVID-19 patients (n=30)**<br>**No. (%)** | **Healthy controls (n=30)**<br>**No. (%)** | ***P*-value** |
| --- | --- | --- | --- |
| IFV-A | 18 (60.00) | 0 (0.00) | **<0.0001** |
| IFV-B | 16 (53.33) | 4 (13.33) | **0.0018** |
| MP | 7 (23.33) | 0 (0.00) | **0.0105** |
| LP | 6 (20.00) | 0 (0.00) | **0.0237** |
| RSV | 0 (0.00) | 0 (0.00) | NA |
| ADV | 0 (0.00) | 0 (0.00) | NA |
| PIV | 0 (0.00) | 0 (0.00) | NA |
| CP | 0 (0.00) | 0 (0.00) | NA |
| COX | 0 (0.00) | 0 (0.00) | NA |
| Total | 24 (80.00) | 4 (13.33) | **<0.0001** |

**Figure 2** showed the rate of coinfection with other respiratory pathogens detected in Qingdao COVID-19 patients as compared with CAP controls. The overall positive rate for coinfection in CAP group was 20.90%, significantly lower than the figure in COVID-19 patients (80.00%, *P*=0.0055). Except for SARS-CoV-2, 7 (23.33%) COVID-19 patients had mixed infections of IFV-A and IFV-B, 6 (20.00%) patients of this group had 3 respiratory pathogens (IFV-A, IFV-B and MP/LP) detected, and 13.33% of them had combined infection of IFV-A only. Whereas the pattern for coinfection in CAP group was quite different: patients were more often coinfected with IFV-B and MP (8.96%), followed by a combination of IFV-A and MP (4.48%), and MP and RSV (2.99%). Thus, it can be seen that COVID-19 patients had distinct aetiological features of coinfection from CAP patients.

**Figure 2.**
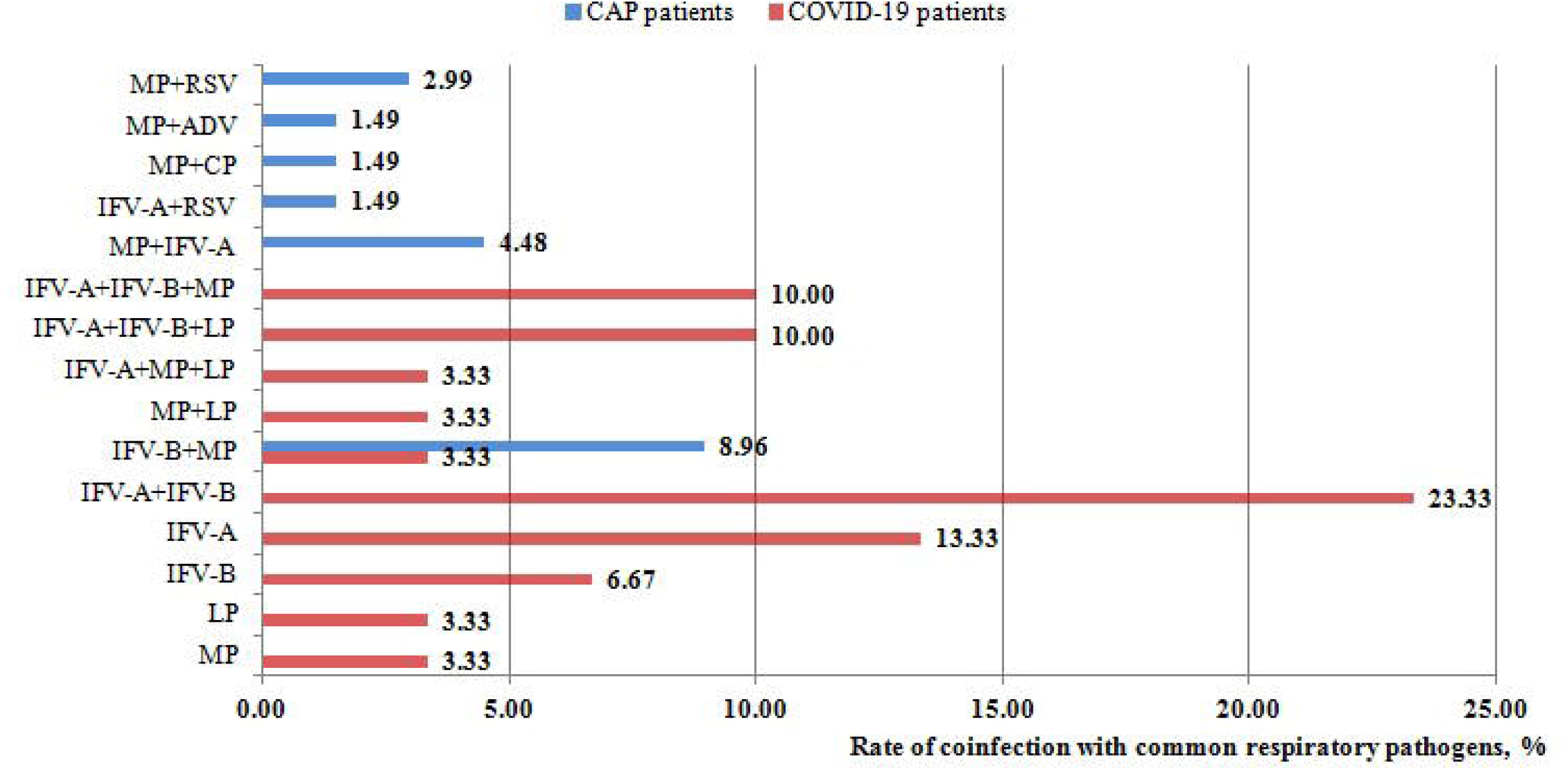
Rate of coinfection with common respiratory pathogens in COVID-19 and CAP patients in Qingdao

The climatic characteristics of Qingdao and Wuhan from December to January of following year were shown in **Table 5**, where differences could be observed between the two cities. Although the climate in Qingdao is drier and colder than Wuhan, major respiratory pathogens circulate in the two cities were quite similar.

**Table 5.** Natural characteristics of Qingdao and Wuhan.

|  | **Qingdao** | **Wuhan** |
| --- | --- | --- |
| Average temperature (°C) | -0.2 - 2.5 | 4.0 - 6.2 |
| Mean humidity (%) | 63 | 73 |
| Epidemic respiratory viruses | IFV-A, IFV-B, RSV, ADV | IFV-A, IFV-B, RSV, and ADV |
| Rate of co-infection (%) | 16.3 | 13.6 |

Average temperature during December and January of following year (Average temperature), Mean humidity during December and January of following year (Mean humidity), influenza virus A (IFV-A), influenza virus B (IFV-B), respiratory syncytial virus (RSV), adenovirus (ADV).

## Discussion

This is an extended descriptive study on COVID-19 patients between Wuhan and Qingdao, represented as within and beyond the epidemic centre, respectively. Since the early onset of COVID-19 till the specific definition of novel coronavirus (previously known as 2019-nCoV) releasing on January 7, 2020,^4^ the term “illness/pneumonia of unknown aetiology” was repeatedly quoted by the health officials and the public during this one-month period. One of the possible explanations for such failure to clearly define the disease is that no causative pathogen could be found at the early stage. We speculated that if any common respiratory pathogen, such as influenza and parainfluenza viruses, RSV, ADV, MP and CP, or the previous emerging novel coronaviruses including SARS-CoV and MERS-CoV, was isolated from body fluids and secretions of the infected patients, related treatment and management could have been implemented in the first place. As such, it was very unlikely that “pneumonia of unknown causes” would still be emphasized, although this might cause delay in the discovery of SARS-CoV-2. Therefore, we assumed that the possibility of coinfection in COVID-19 patients was very rare. Our assumption was led support by two recent studies conducted in Wuhan showing that there was no coinfection of respiratory pathogens in COVID-19 cases.^3,9^

According to our data, the majority of COVID-19 patients in Qingdao were not from the endemic area; they were infected indirectly without a history of traveling to Wuhan. Despite the high coinfection rate in Qingdao patients who had more complicated clinical conditions (who were elder with more underlying diseases), there are no signs of worsening of these patients’ clinical manifestations or disease prognosis as compared with Wuhan patients. Whether immune regulation associated with earlier infection with common respiratory pathogens confers an element of protection against the pathological damage caused by subsequent infection of SARS-CoV-2 remains unanswered. A study conducted in Zhejiang Province, China revealed that COVID-19 patients outside of Wuhan had relatively mild symptoms as compared with those initially reported in Wuhan.^13^ Like patients in Qingdao, these Zhejiang patients had no direct contact with the original site of outbreak (Wuhan Huanan Seafood Market). Evidence is scarce at the present stage to explain whether it is due to the weakening of SARS-CoV-2 virulence during virus transmission. Previous studies on SARS have shown that the disease was more infectious at an earlier stage when compared to the later stage.^14^ The global mortality rate of MERS was about 40% and declined sharply to less than 20% during second generation transmission.^15^ Currently, our first priority is to slow transmission of SARS-CoV-2. Some of the patients are still under hospitalised treatment or in quarantine, requiring a multidisciplinary approach to illustrate the underlying mechanisms in the future.

Here we also compared the geographical and climatic characteristics of Qingdao and Wuhan. Wuhan is located in the centre of southern China and has a subtropical climate;^16,17^ while Qingdao is situated in the coastal area of northern China in the temperate zone, which has a relatively lower humidity than Wuhan.^18^ Despite the difference in natural characteristics between the two cities, common respiratory pathogens circulate in Wuhan and Qingdao (including IFV-A, IFV-B, RSV, and ADV) have shown to be generally similar during the peak season of respiratory diseases in wintertime (from January to February).^19-22^ The incidence of coinfection in COVID-19 patients in Wuhan was rather low. Whereas in places with relatively low temperature like northern Qingdao, it is more common to find combined SARS-CoV-2 infection with other seasonal respiratory pathogens. It is not yet known whether this phenomenon also exists in other regions, leaving a gap for future studies. There are limited data to address whether coinfection with other respiratory pathogens would affect the pathogenesis and outcome of severe acute respiratory illnesses like SARS and MERS.^23-27^ Fortunately, SARS-CoV-2 RNA was detected in all of our patients before other respiratory pathogens being detected. Otherwise these COVID-19 patients would have been treated in a way as infected with other respiratory pathogens, which might result in devastating consequences. Our findings provide some important implications for the prevention and management of COVID. First of all, it is very effective and efficient for the China CDC and local governments to launch series of aggressive measures to screen, quarantine, diagnose, treat and monitor suspected patients and their close contacts in response to the widespread transmission of the virus. Moreover, we must remain vigilant when dealing with COVID-19 cases in other locations outside of Wuhan. Even common respiratory pathogens are detected by diagnostic testing, we still need to rule out the possibility of SARS-CoV-2 infection. In light of the emergence of coinfected COVID-19 cases, there might be a long-term coexistence of SARS-CoV-2 with humans, just as other seasonal respiratory pathogens. Last but not least, we highly recommend adding SARS-CoV-2 to the diagnostic testing assay as soon as possible, in a manner as how we routinely screen influenza and parainfluenza viruses, RSV, ADV, MP and CP, etc. Our hospitals adopted the above-mentioned screening and surveillance protocol right after the recognition of first coinfected COVID-19 case. Further detailed investigations should aim to ascertain the exact reasons underlying the high coinfection rate outside of Wuhan.

It is notable that there are several limitations of this study. Two critically ill patients and one patient who died in the end were excluded due to lack of aetiological data other than SARS-CoV-2. As of February 16, 2020, there were 60 cases of confirmed SARS-CoV-2 infection in Qingdao, however, we only analysed 30 non-severe cases whose complete aetiological and clinical information was available. The age of patients in Qingdao ranged from 1.5 years to 80 years; whereas patients in Wuhan were all adults as they were medical professional infected through work. Such age difference between the two groups might bring in bias to this study. We performed nucleic acid testing for the confirmation of SARS-CoV-2 infection, while coinfection with other pathogens was detected by serological testing of antibodies. In spite of superior accuracy, PCR-based molecular testing using throat swabs requires more complicated techniques of laboratory personnel with prolonged reporting time, which may not be suitable for emerging cases of SARS-CoV-2.^28,29^ Thus, we applied rapid testing of aetiological agents by IIF in collected acute phase serum to guide clinical decision making. Since both the hospitals in Qingdao and Wuhan adopted the same protocol for the diagnosis of coinfection, we believe this will not affect the determination of final outcomes.

To test the reliability of serum specific IgM detection, we recruited a control group of normal people without clinical symptoms. The low infection rate in our control population suggested that our method was reliable in early and rapid diagnosis of respiratory infections. However, we were unable to exclude the possibility that coinfection with other respiratory pathogens may make the patients more susceptible to SARS-CoV-2 infection. Specific-IgM antibody could be detected in serum from one week of illness onwards and the amount progressively increased and peaked at about two weeks after infection.^30^ Positive results for serum IgM antibody detection only indicate infection with specific pathogens during the past one to two months but cannot specify the time sequence of coinfection with various pathogens.

Till now, the epidemic is still expanding with the number of infected patients escalating rapidly, and the situation in countries and regions beyond China presents a more dismal picture. Recently, WHO upgraded the risk assessment of SARS-CoV-2 to very high; as of March 1, 2020, a total of 7,169 cases in 58 countries, and 104 deaths have been reported outside China.^8^ Hence, we call for a timely action to carry out real-time research on the epidemic dynamic of SARS-CoV-2.

## Data Availability

The complete dataset is included in this manuscript

## Contributor

Q-SX and Y-HX designed the study and conceptualised the paper. TC, KD, R-QG, C-ZC, YG, QL, G-LY, J-NT, YS, AZ, QW, G-RM, J-WC, Z-PL and S-LP collected the epidemiological and clinical data. W-JL, WL, GL-H contributed to laboratory testing. G-JL and WN summarised the data and conducted statistical analysis. Q-SX, Y-HX, G-JL and WN wrote the initial draft of the manuscript. All authors provided critical feedback and approved the final version. The corresponding author has full access to all data in this study and attests that all listed authors meet authorship criteria and that no others meeting the criteria have been omitted.

## Data sharing

After publication, the data that support the findings of this study will be made available from the corresponding author on reasonable request. It will be necessary to provide a proposal with detailed description of study objectives and statistical analysis plan for evaluation of the reasonability of requests. Additional materials might also be needed during the process of evaluation. Deidentified participant data will be provided after approval from the corresponding author and Qingdao Municipal Centre for Disease Control and Prevention, Qingdao Women and Children’s Hospital, Affiliated Hospital of Qingdao University Medical College, Qingdao Chest Hospital and Renmin Hospital of Wuhan University.

## Acknowledgements

This study is supported by The National Natural Science Foundation of China (NSFC) [Grant number 81770315]; and Distinguished Taishan Scholars (2019). We thank Municipal Centre of Disease Control and Prevention of Qingdao for coordinating data collection for COVID-19 patients in Qingdao. We are deeply thankful to all health-care workers involved in the diagnosis and treatment of patients in Qingdao and Wuhan. And we thank Prof Gary WK Wong for guidance in study design and interpretation of results.

